# Disagapp: A Shiny app to facilitate reproducible disaggregation regression analyses

**DOI:** 10.64898/2026.09.09.26362604

**Authors:** Simon E. H. Smart, Olukemi O. Olowofoyeku, Tim C. D. Lucas

## Abstract

Creating high-resolution maps of disease risk is important for many climatically-driven diseases including vector-borne, zoonotic and non-communicable diseases. However, disease counts are commonly only available aggregated to an administrative level such as the county, department or province. Using disease mapping to make highresolution risk maps from these data can be challenging, even though high-resolution data on temperature and other environmental variables are available. Disaggregation regression is a new multiscale modelling framework that has been used in mapping global malaria incidence and dengue incidence, amongst others. We present Disagapp (https://github.com/simon-smart88/disagapp; https://disagapp.le.ac.uk/) a web app for disease mapping with administrative-level data. The app was designed to be user-friendly, with a point and click interface and with modelling guidance integrated at each step of the analysis. It can be accessed online or run locally by installing the R package and calling one function. Environmental and economic covariates (temperature, precipitation, distance to water, land use, population density, accessibility and night lights) are seamlessly retrieved from various sources, collated and harmonised and used to fit disaggregation regression models using the disaggregation R package. This modelling framework is a principled way to create high-resolution predictions of disease risk. Analyses conducted in the app can be reproduced outside of the app by generating an R markdown document, allowing the user to share, preserve, extend or learn from their analysis. By making the techniques available in the disaggregation package available online and making it simple and easy to access covariates, we remove barriers for non- or novice R users or analysts who work in institutions with restrictive installation permissions to access these innovations. Therefore, this new app makes this important modelling framework much more available to the global modelling community.

## 3 Background

### 3.1 Disease mapping

Creating high-resolution maps of disease risk is important for many climatically-driven diseases including vector-borne, zoonotic and non-communicable diseases (Lim et al, 2025; Weiss et al, 2025; Reiner Jr and Hay, 2022; Zhao et al, 2024). High resolution maps enable more targeted interventions, which are critical to meeting Sustainable Development Goals (Utazi et al, 2019). However, disease counts are commonly only available aggregated to an administrative level such as the county, department or province. Using disease mapping to make high-resolution risk maps from these data can be challenging, even though high-resolution data on temperature and many other environmental variables are freely available from remote-sensing and other sources (Weiss et al, 2018; Fick and Hijmans, 2017).

### 3.2 Disaggregation regression

Disaggregation regression is a method for generating high-resolution predictions from response data aggregated over large areas (e.g. counties or states) by using covariates to inform the heterogeneity within polygons. It was first applied as a method for modelling species distributions (Keil et al, 2013) but has found wider usage in epidemiology in order to generate high-resolution maps of disease risk. The R package *disaggregation* (Nandi et al, 2023) implements methods for disaggregation regression using TMB or template model builder (Kristensen et al, 2016). The disaggregation package has been applied in epidemiology (Lucas et al, 2022; Nightingale et al, 2024; Poongavanan et al, 2025), ecology (Murphy et al, 2023), environmental management (Clark et al, 2025) and economics (Caset et al, 2023). Models fitted with *disaggregation* are Bayesian disaggregation regression models, with optional spatial random effects and individually and identically distributed (IID) random effects to account for overdispersion of counts (Nandi et al, 2023). The overarching aim of these models is to connect high-resolution covariates with low-resolution response data, aggregated to some set of geographic polygons such as administrative boundaries.

Several forms of data are required to fit a disaggregation regression model, making it a complex task which requires expertise in manipulating spatial data in R. The response data is the areal data for which we wish to fit a model to; polygons define the spatial coordinates that each row of the response data corresponds to; covariates are the high resolution data used to predict the response data; the aggregation data, typically population counts, informs the model of where cases could occur inside each polygon; a spatial mesh is required for the computational approximations used by *disaggregation* to speed up model fitting.

Disagapp aims to make the process of fitting disaggregation models easier and to increase the use of the method by making it available to analysts who do not have sufficient experience of programming in R to use the disaggregation package, or who work in organisations with restrictive IT permissions that may prevent installation of software. The app also has advantages to more sophisticated users, by providing easy access to a wide range of covariates and by generating code for model fitting that can then be extended as required.

### 3.3 Shiny apps

Shiny is an R package used to develop interactive apps that can either be run locally or deployed to web servers (Chang et al, 2024). It is an increasingly popular method for researchers to develop apps due to the relative ease of development compared to other approaches (Kasprzak et al, 2021). Developing and deploying apps enables users to access the analytical power of R from a web browser, without the requirement of installing all of the software dependencies. Conducting analyses in an app can have disadvantages however as it may be difficult to reproduce the analysis in the future, either because the precise settings used cannot be recalled, the app is no longer accessible or because dependent software has been updated resulting in breaking changes. Thus, in developing Disagapp we aimed to develop an easy-to-use interface for conducting the analysis, but also generate a downloadable R script which can be run to reproduce the analysis.

### 3.4 Paper structure

In Section 2 we discuss how the app was designed and implemented, detailing how the app is structured and it’s general features. In Section 3 we provide a walkthrough of the app detailing how a user can perform a complete analysis.

## 4 Implementation

Disagapp was created with *shinyscholar* (Smart and Lucas, 2025), a template for creating reproducible analytical Shiny apps developed by forking *WallaceEcoMod* (Kass et al, 2023). It is structured as an R package, facilitating distribution and installation.

### 4.4 Structure and layout

The app is structured into *components* representing stages in the analysis and *modules* representing possible options at each stage. There are seven components; *Response, Covariates* and *Aggregation* each load their respective types of data. Aggregation data is a raster layer, often of population counts, used as an offset in a manner similar to in Poisson GLMs. *Prepare* modifies the data into a consistent format. *Fit* fits the model. *Predict* generates predictions and *Reproduce* enables the analysis to be reproduced outside of the app. Modules are colour-coded depending on whether they are mandatory to use (orange), at least one of the modules in the component must be used (blue) or optional (pink).

Components are navigated between using the bar at the top of the app. Within each component, a menu lists the modules available and selecting one loads the options for that module. Modules are run by pressing the button at the bottom of the options menu and outputs are displayed in the Map and Results tabs. Guidance on how to use each component and module is available inside the results panel. Apart from the map, the contents of the tabs in the results panel depend on the module currently selected. If any errors occur, they appear as a pop-up and are also included in the log window (Figure 1).

**Fig. 1:**
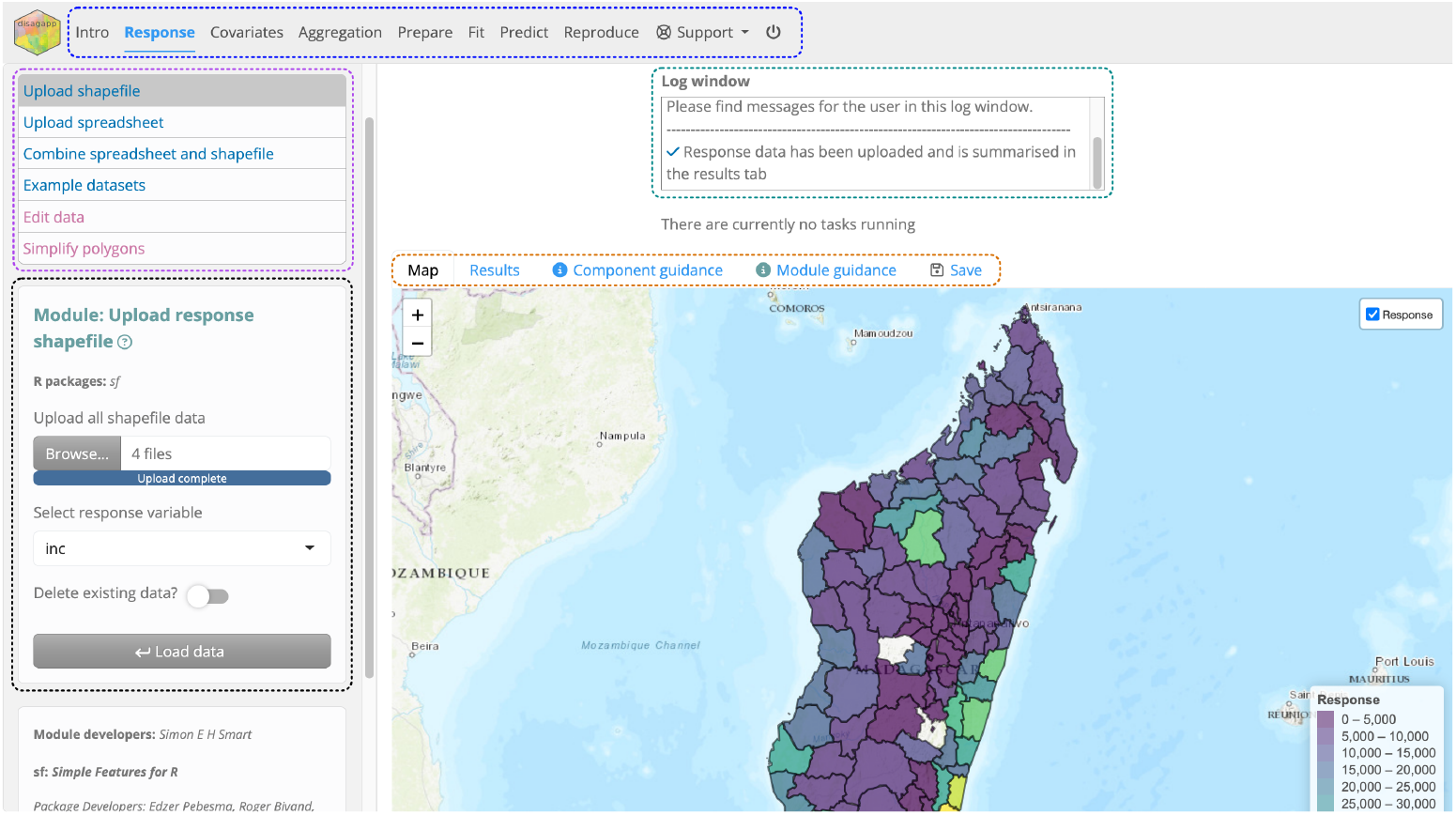
Annotated screenshot of the app when the *Upload shapefile* module is in use showing the navigation bar for switching between components (blue), the menu for switching between modules (purple), the options for the selected module (black), the log window (teal) and the results panel (orange).

### 4.2 Development and general features

The layout and structure visible to the user is replicated in the app architecture with each module having four associated files located in `inst/shiny/modules`. Most modules have one synonymous function located in `R/` although some are shared between modules (e.g. `cov_landuse` and `agg_landuse`) or call functions from the disaggregation package instead. Each module consists of a Shiny module, an Rmarkdown document that reproduces the module, a markdown document containing guidance on how to use the module and a configuration file detailing the component that it belongs to and the packages used to create it. The Shiny modules are composed of a user interface function that generates the options visible in the sidebar, a server function where the computation occurs, a results function which generates outputs, a mapping function that updates the map and an rmd function which passes parameters to the module’s Rmarkdown document.

The general features of Disagapp are inherited from shinyscholar. These include a logger where messages to the user are presented, the ability to save and load the analysis at any point, an interactive map built with *leaflet* (Cheng et al, 2024), an introductory walkthrough powered by *rintrojs* (Ganz, 2016) and the ability to add new modules when the app is run locally. A logging window is included in the user interface to which messages can be sent from either inside module functions or within the module itself. These messages can just provide information to the user or they can be error messages that prevent further evaluation of a function. Slow-running operations, for example downloading covariates and fitting the model, are run asynchronously using `shiny::ExtendedTask()`, *future* (Bengtsson, 2021) and *promises* (Cheng, 2024) which enables the app to remain responsive whilst these operations are in progress and for multiple operations to occur in parallel. Each component has an associated .Rmd file that provides guidance on the modules within it and each module has an .md file that provides further details on the module, including instructions on how to use it, the theoretical basis and relevant citations to the literature. Overall, this guidance aims to provide all the information that the user requires to complete the analysis, so that no further materials, such as user guides, videos or instruction manuals are required. Packages used to develop each module are cited within the user interface and links to their documentation are provided.

Data is passed between modules through an R6 object named `common` which is defined in `inst/shiny/common.R`. Objects within `common` approximately correspond to the components. For example, the *Response* modules create or edit `common`$`shape` and covariates are initially stored in `common`$`covs` and then `common`$`covs prep` once they have been prepared for analysis. The `common` object also contains utility objects including: the logger; `meta` where module inputs are stored when they are run; `state` where module inputs are stored when the app is saved; `tasks` where long-running tasks are stored. The `common` object is converted to a list when the app is saved and the opposite operation occurs when the app is loaded. `common`$`reset()` resets the data structure and is called when a new dataset is loaded in the *Response* component.

Development was tracked using git for version control and stored online via GitHub at https://github.com/simon-smart88/disagapp. Unit tests for module functions were written using *testthat* (Wickham, 2011) and end-to-end tests for individual modules and overall analyses were written using *shinytest2* (Schloerke, 2024). Continuous integration was implemented using GitHub Actions to both run `R CMD check`, and the tests, on multiple operating systems whenever changes were pushed to the repository (Wickham and Bryan, 2023). A detailed guide to developing new modules is included in the repository in the `adding_modules.md` document.

### 4.3 Components and modules

Information on each module in Disagapp is provided in Table 1. In addition to the user-facing Components previously described, four `core` modules provide app functionality that is separate to the analysis (mapping, saving, loading, introductory tour) and so only consist of a Shiny module and are not reproducible.

**Table 1:**
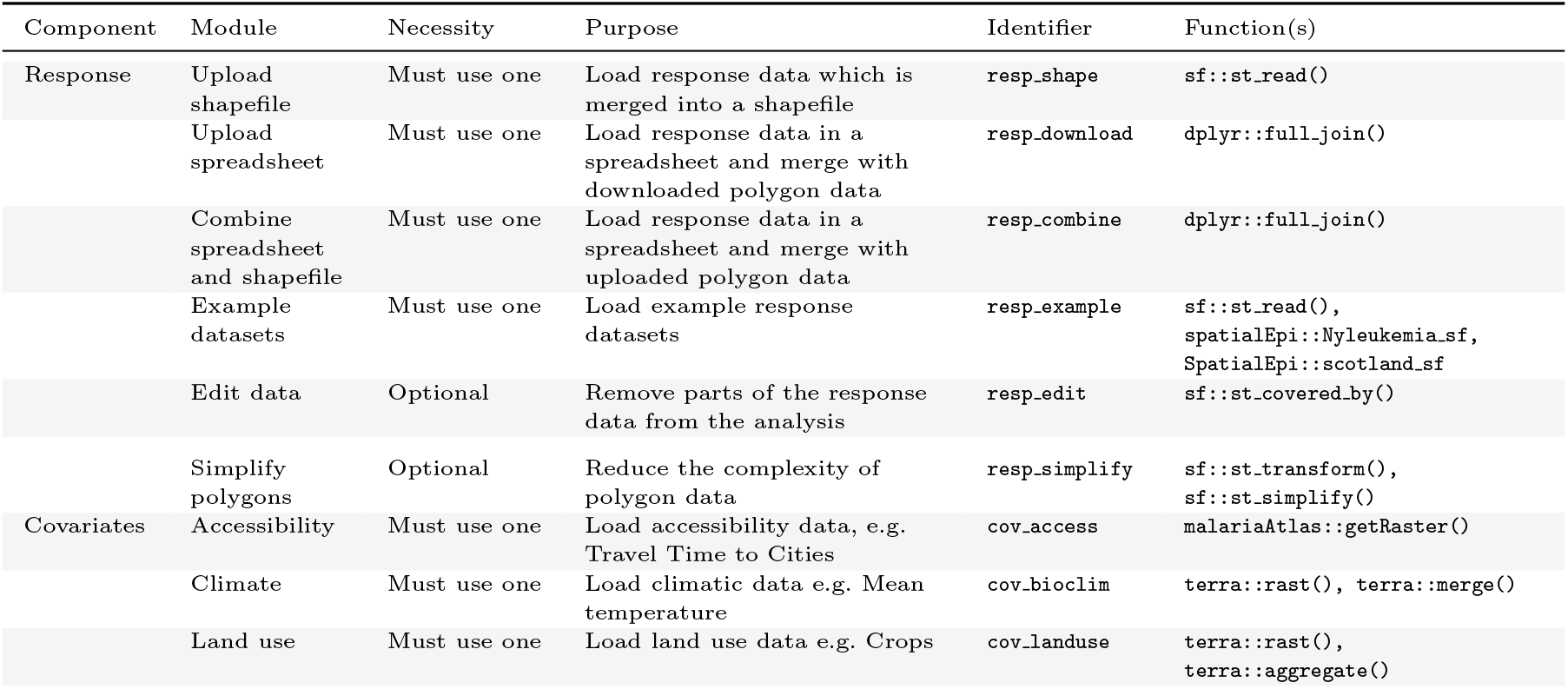
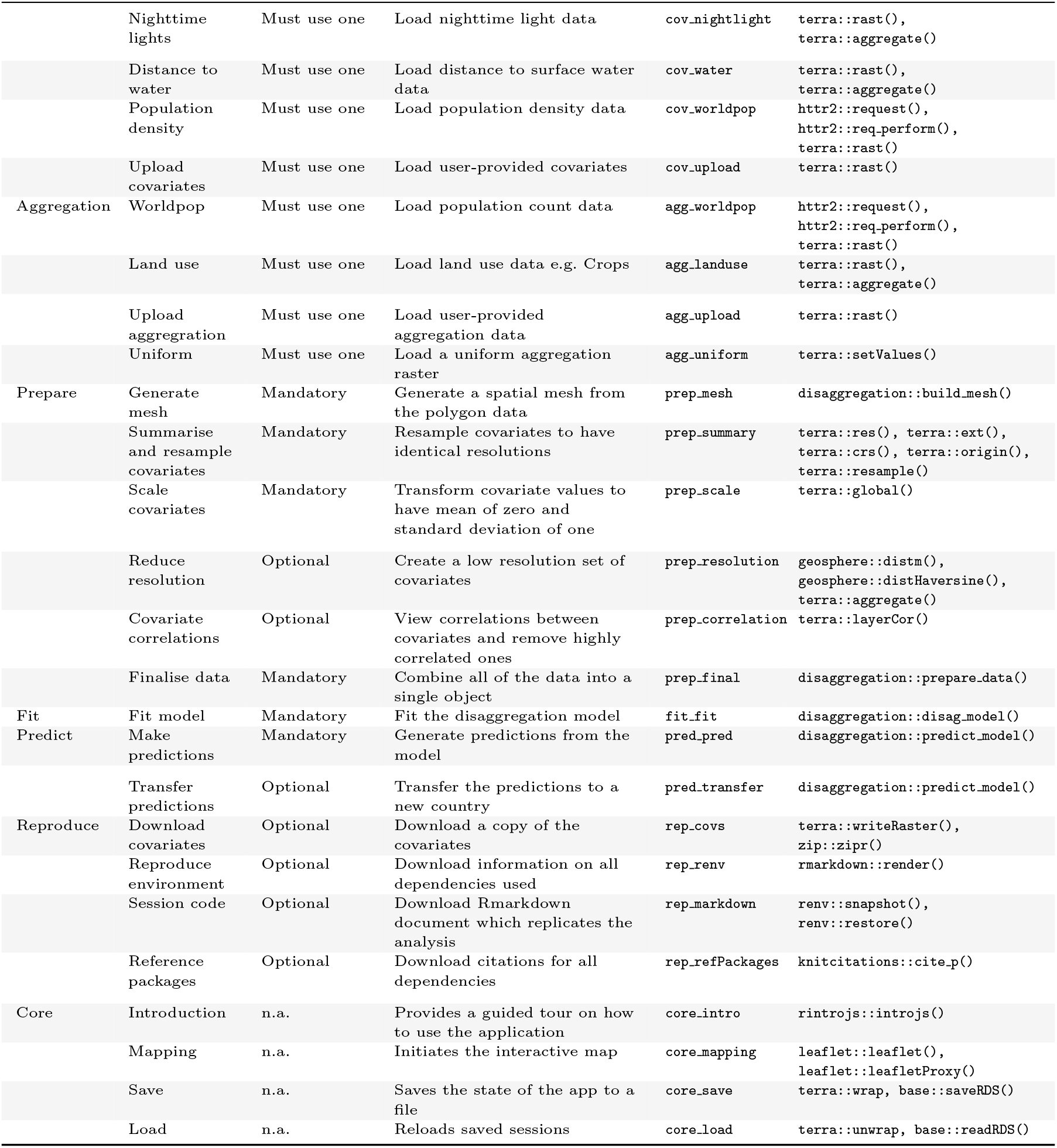
Components and modules in Disagapp. For each module the necessity of using the module, the purpose of the module, the identifier which is used to create files and module functions and the dependent functions are noted. The functions are named using the syntax of R package::function. n.a. = not applicable

### 4.4 Mapping

The base map is created in the `core mapping` module using *leaflet* and this module returns a `leafletProxy` object named `map` which is passed to each module to enable them to interact with the map. Several functions facilitate plotting of objects on the map in a consistent manner i.e. with a suitable name, a legend and being included in the panel for selecting visible layers. `Shape_map()` plots the response data, `raster_map()` plots raster data and `mesh_map()` plots the spatial mesh. Once the covariates have been prepared and if the low resolution set of covariates have been created, users can switch between viewing the original covariates, prepared covariates or low resolution covariates using a menu below the map which triggers `replot_raster_map()` to be called and load the selected data to the map.

### 4.5 Reproducibility

Computational reproducibility is an important principle in all scientific analyses and Disagapp enables any analyses completed in the app to be fully replicated outside of it by creating a downloadable Rmarkdown document. Each module has an associated Rmarkdown file that calls the same function as the module. When the module is run, a flag is recorded in `common`$`meta` alongside all of the input settings when the module is run. When the *Download session code* module is run each module’s `rmd_function` is called and the input values are *knitted* into the module’s Rmarkdown document only if the relevant module has been run by the user. The individual Rmarkdown chunks are then combined together to create a single document which if rerun in R will produce the same outputs. One limitation is that it is not possible to determine the location of uploaded files and thus, the Rmarkdown document only contains the file name and the user must add a path to where the file is located.

Updates to dependent packages could affect the ability to reproduce the analysis in the future. To address this, the *Reproduce environment* module captures the versions of all dependent packages using `renv::snapshot()` to produce a downloadable .lock file. Once the module is run the Rmarkdown is updated to begin by reading the lock file and install the exact same package versions using `renv::restore()`.

### 4.6 Deployment

Deployment, where the app is run on a web server and therefore available to any user with a web browser was a crucial requirement for the aims of disagapp to be easily available to analysts. The app is available at https://disagapp.le.ac.uk and is run on Shiny Server. The server has 16 GB of RAM and 8 cores enabling analyses to be performed that contain over 10^6^ cells and enabling multiple asynchronous tasks to run at the same time.

## 5 Results

This section provides a guide for a simple but complete analysis in the app. A variety of additional options, beyond those described in this walkthrough, are available (Table 1). Different methods for reading in data and downloading covariate data have been implemented and additional modules for exploratory analysis are also available. Details and the full range of options have been deliberately avoided as these are documented in detail in the app itself.

### 5.1 Loading response data

In this example we will load data using the *Upload shapefile* module which requires that the response data and polygon data for the administrative areas are already combined. Shapefiles are composed of four separate files and the first step is to upload these by clicking on the *Browse* button and navigating to the location of the files. Select the files and press Open to upload them to the app. Once loaded, the *Select response variable* option becomes visible and this allows selecting the column in the data containing the response data to analyse. Click the dropdown menu and select the relevant column, in this case *inc*, and then click *Load data*. After a few moments, the data will be displayed on the interactive map and also summarised in the Results tab (Figure 1). The data used in this example is also available through the *Example datasets* module by selecting the default *Malaria in Madagascar* dataset.

### 5.2 Loading covariate and aggregation data

Once the response data has been loaded, we can load the covariate data. In this example we will use the *Climate* module which downloads the user’s selection of 19 covariates summarising temperature and precipitation (Fick and Hijmans, 2017). For this module, the data is initially downloaded for separate countries, so the first step is to select the name of the country from the list. The list can be searched by typing in the first letters of a country and if the response data covers multiple countries, these should all be included. Whichever countries are selected are automatically used in other covariate modules. Next, select the Bioclim layers of interest from the menu and then click *Load data*. Unlike when loading the response data, after pressing this button it will display *Processing* and the notice below the log window will update to say that one task is in progress, informing the user that the data will be loading in the background and that they can continue using the app. Once this is displayed, we can proceed to load more covariate data using other modules, When the data has been downloaded, it will be displayed on the map and summarised in the Results tab. To switch between the layer displayed on the map, use the layer selection panel in the top right-hand corner of the map (Figure 2). Loading aggregation data is similar to loading covariate data and for epidemiological use cases, the default values in the *Worldpop* module are suitable and so clicking *Load data* is sufficient.

**Fig. 2:**
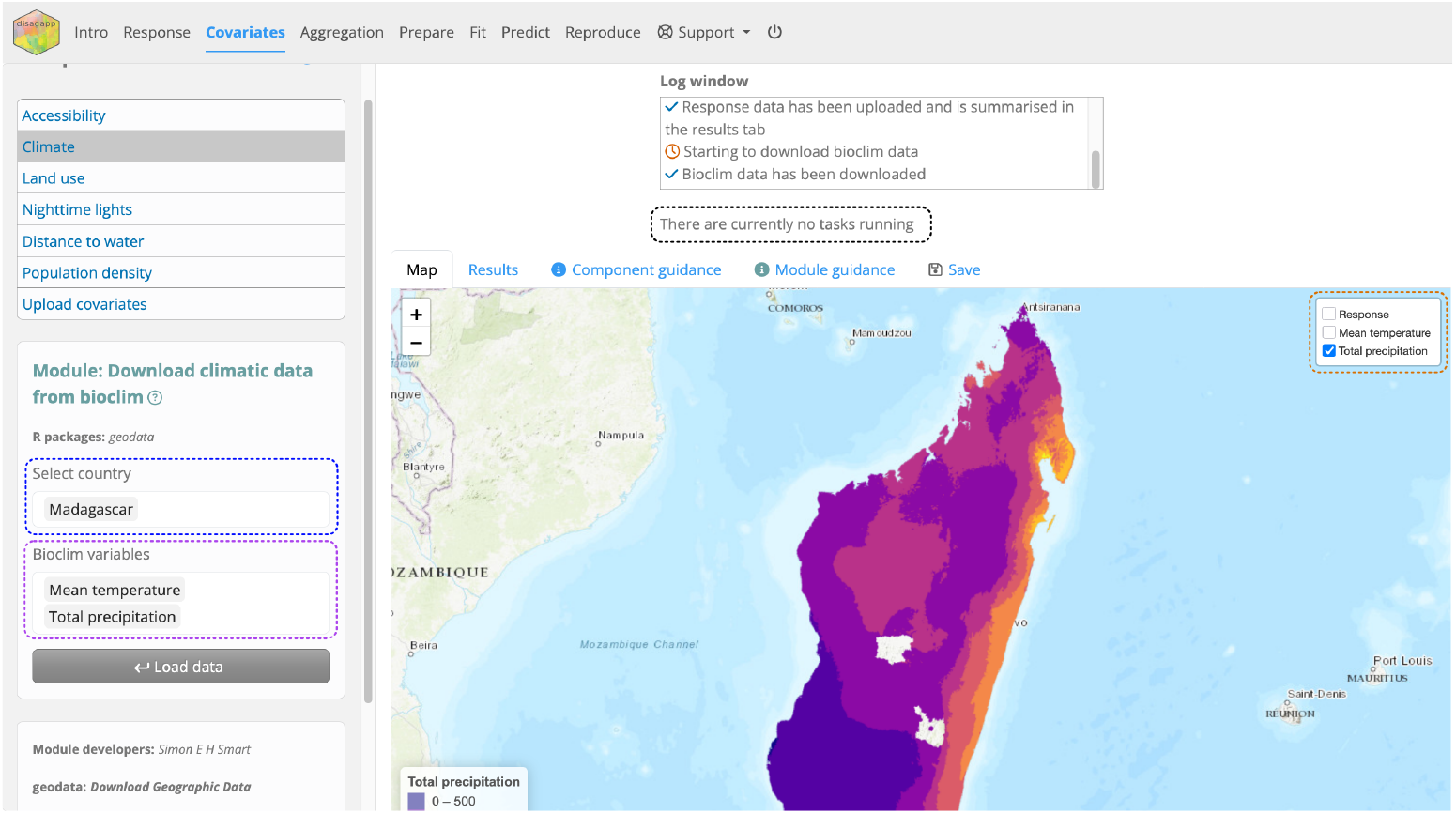
Annotated screenshot of the app when the *Climate* module is in use, showing the country selection input (blue), the Bioclim variable selection input (purple), the background task status (black) and the map layer selection panel (orange).

### 5.3 Preparing data

Unlike in the previous components where either a single or multiple module could be used depending on the analysis, in the *Prepare* component there are four modules which must be used. *Generate mesh* produces a spatial mesh. This is a data object that is needed for computational approximations for the spatial random effect (Nandi et al, 2023; Bakka et al, 2018). The default values are typically suitable for an initial model run. For final model runs, it is advisable to generate a denser mesh containing 2000-3000 nodes. As a general principle it is good modelling workflow to limit the run time of models during model development, and to then increase back to full data precision of resolution for final model runs. The module guidance contains detailed information on how the parameters can be adjusted to produce different meshes and multiple meshes can be produced which are displayed in the Results tab. Once meshes have been generated, the mesh to use in the analysis can be selected in the dropdown menu (Figure 3).

**Fig. 3:**
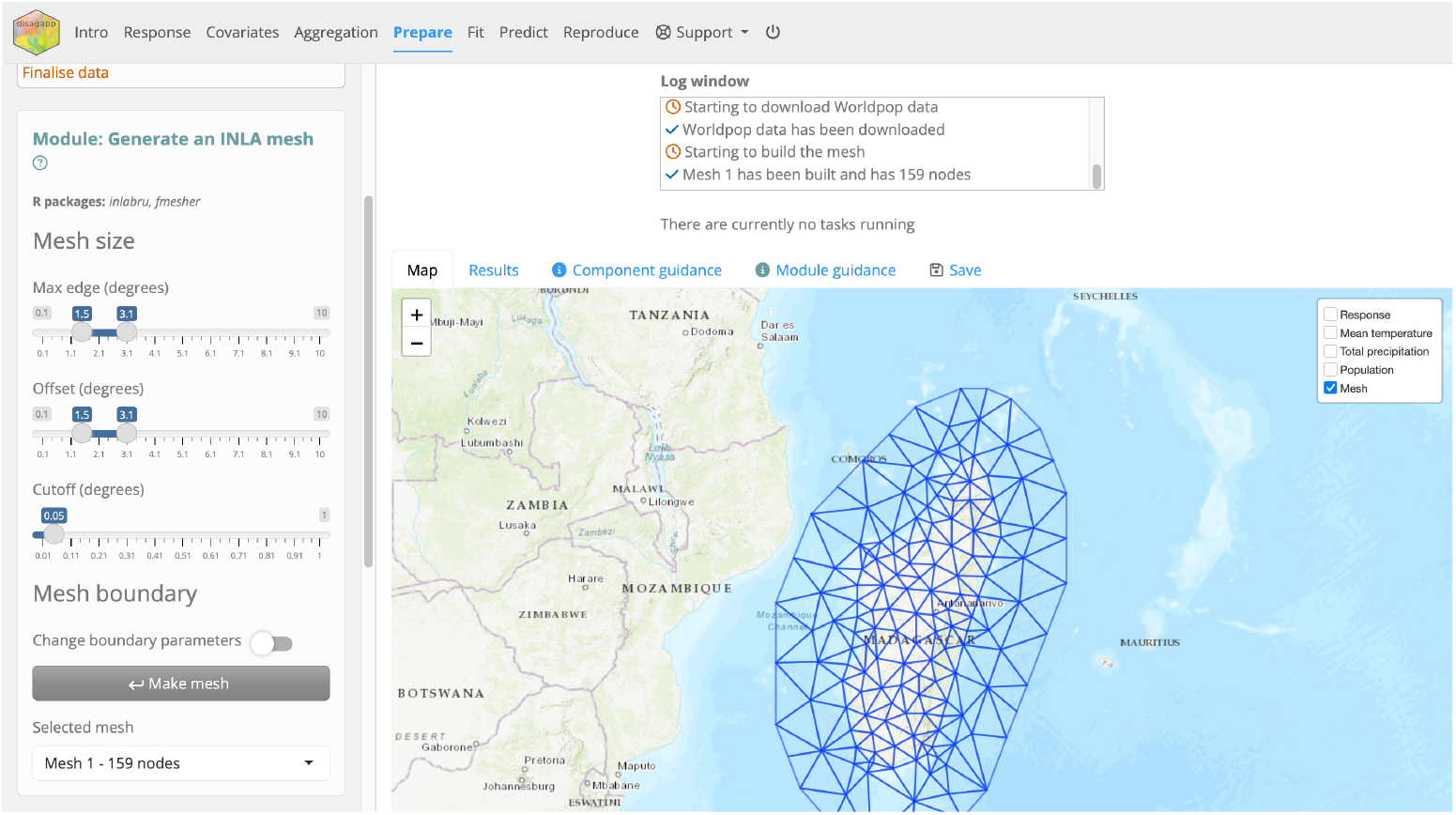
Screenshot of the app when the *Generate mesh* module has been run.

The covariate and aggregation data must be made consistent with each other by resampling so that each raster has the same resolution and origin, i.e. each cell aligns perfectly with the cells in the other rasters. Click *Prepare covariate summary* in the *Summarise and resample covariates* module to generate a summary table of the rasters detailing their resolution. The *Remove identical rows?* switch can be toggled to only show properties which differ. Once the table is generated, a dropdown menu becomes visible and one of the rasters should be chosen to use as a template to resample the other rasters. Click *Resample covariates* and the resampled covariates will become visible on the map. The original rasters can be viewed again, by toggling the *Covariates* menu which appears below the map. The original covariates can have highly contrasting ranges in values, for example total precipitation in Madagascar (measured in millimetres) ranges from 0 to 3500 whereas mean temperature (measured in degrees Celsius) ranges from 12 to 28. These differences would make it difficult to interpret the coefficients of the model but this can be resolved by scaling the covariates so that they all have mean of zero and a standard deviation of one by clicking *Scale covariates* in the *Scale covariates* module.

The *Reduce resolution* module produces a set of covariates with lower resolutions, reducing the time required to fit a model. The module is optional, but it is generally advisable to use for initial runs since the covariates typically have a resolution of *c*. 1000 m which is excessive during model development. Click *Summarise data* to produce either a histogram or boxplot showing the distribution of cells per polygon at the original resolution. Choose a new resolution from the *New cell width* dropdown menu; the available options are all multiples of the original resolution and typically a resolution of 5000 m is suitable (which reduces the number of cells by a factor of 25). Click *Reduce resolution* and the lower resolution covariates will be visible on the map. The original resolution covariates can be restored using the menu beneath the map.

The *Finalise data* module collates all the data into a single object, ready to fit the model. From the *Select ID variable* menu, choose the column in the uploaded shapefile which contains a unique identifier for each polygon. All the data can be viewed by returning to the *Upload shapefile* module and viewing the Results tab. If a set of low resolution covariates has been generated, choose to use these or the original high resolution covariates. The *Handle missing data?* switch handles missing covariate and response data in different ways. While these might be appropriate for initial runs, more careful thought should be given for final model runs. Finally, click *Prepare data*.

### 5.4 Fitting the model and generating predictions

The *Fit* component contains a single module for fitting the model. In most epidemiological use cases, the default options are suitable for fitting the model and the module guidance explains when it may be suitable to alter the defaults. The model typically contains covariates, a spatial random effect (a smooth spatial surface that can be thought of as accounting for missing covariates) and an IID effect. This IID effect can be thought of as modelling overdispersion as the assumed Poisson likelihood is making strict assumptions that are often not met in real-world data. This IID approach can be thought of as an alternative to using a negative binomial likelihood. Click *Fit model* and the model fitting will begin. Depending on the number of cells in the covariates and the number of nodes in the mesh, this may take several minutes to run while larger models may take an hour or more. Once complete, the Results tab displays three plots; the first two show the model parameters with the left panel displaying the hyperparameters of the random effects and intercept and the right panel showing a parameter for each covariate; the lower panel shows a scatter plot of the observed and predicted values for each polygon, both including and excluding the IID effect of the model (Figure 4). The IID effect is very flexible and allows the predictions to almost exactly match the observed data, but this is not indicative of the true predictive performance of the model, which is better represented by the predictions made without the IID effect.

**Fig. 4:**
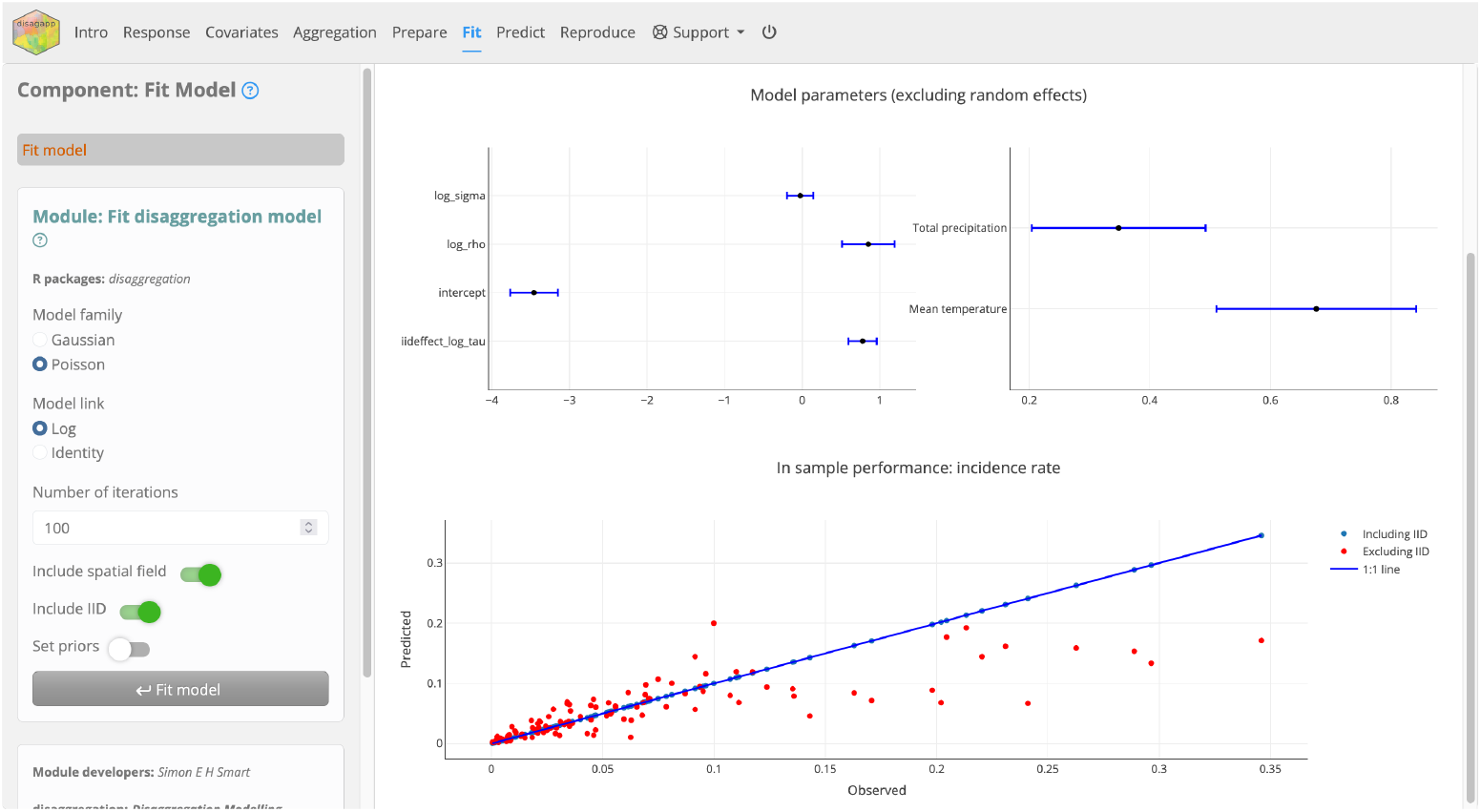
Screenshot of the app after the *Fit model* module has been used.

The *Predict* component uses the fitted model to generate predictions. The *Make predictions* module applies the model to the region of interest defined by the response data to produce a high resolution prediction. By default, the module produces a raster of the predicted rate excluding the IID effect. Switches can be toggled to instead generate predictions that include the IID effect, also produce a raster displaying the rate converted to cases and rasters quantifying the uncertainty of the prediction. When the *Include uncertainty?* switch is toggled, options appear to select the number of realisations and the desired credible interval to summarise the uncertainty. Click *Produce predictions* to run the module and the selected predictions will be added to the map. Once the predictions are generated, the *Download model predictions* button becomes visible and clicking this downloads the predictions as `.tif` files. The *Transfer predictions* module enables transferring predictions to another country. This requires downloading the covariate data for the new country which occurs automatically but can take a long time depending on the number of covariates and the size of the selected country.

### 5.5 Reproducing the analysis

The *Reproduce* component is optional to use but contains modules which ensure the long term reproducibility of the analysis. Due to the covariates originating from various online sources, they may become unavailable in the future. The *Download covariates* module allows downloading a copy of the original (unprocessed) covariates as a `.zip` file. Updates to R packages used in the app could break the existing version and prevent the same code producing the same predictions. To negate this, the *Reproduce environment* module produces a `.lock` file containing information on each R package installed on the server which can be used to reinstall the exact same versions on another computer. The *Session code* module produces an Rmarkdown document which can be rerun to reproduce the analysis conducted in the app (Figure 5). If the *Download covariates* or *Reproduce environment* modules have been used, the Rmarkdown document is modified to read in the `.zip` and `.lock` files so that the covariates are not downloaded again and the R package versions are identical to in the original analysis. If these modules have not been used, the Rmarkdown file contains the code to download the covariates. Finally, the *Reference packages* module produces a list of citations to R packages used in the app, suitable for inclusion in a manuscript.

**Fig. 5:**
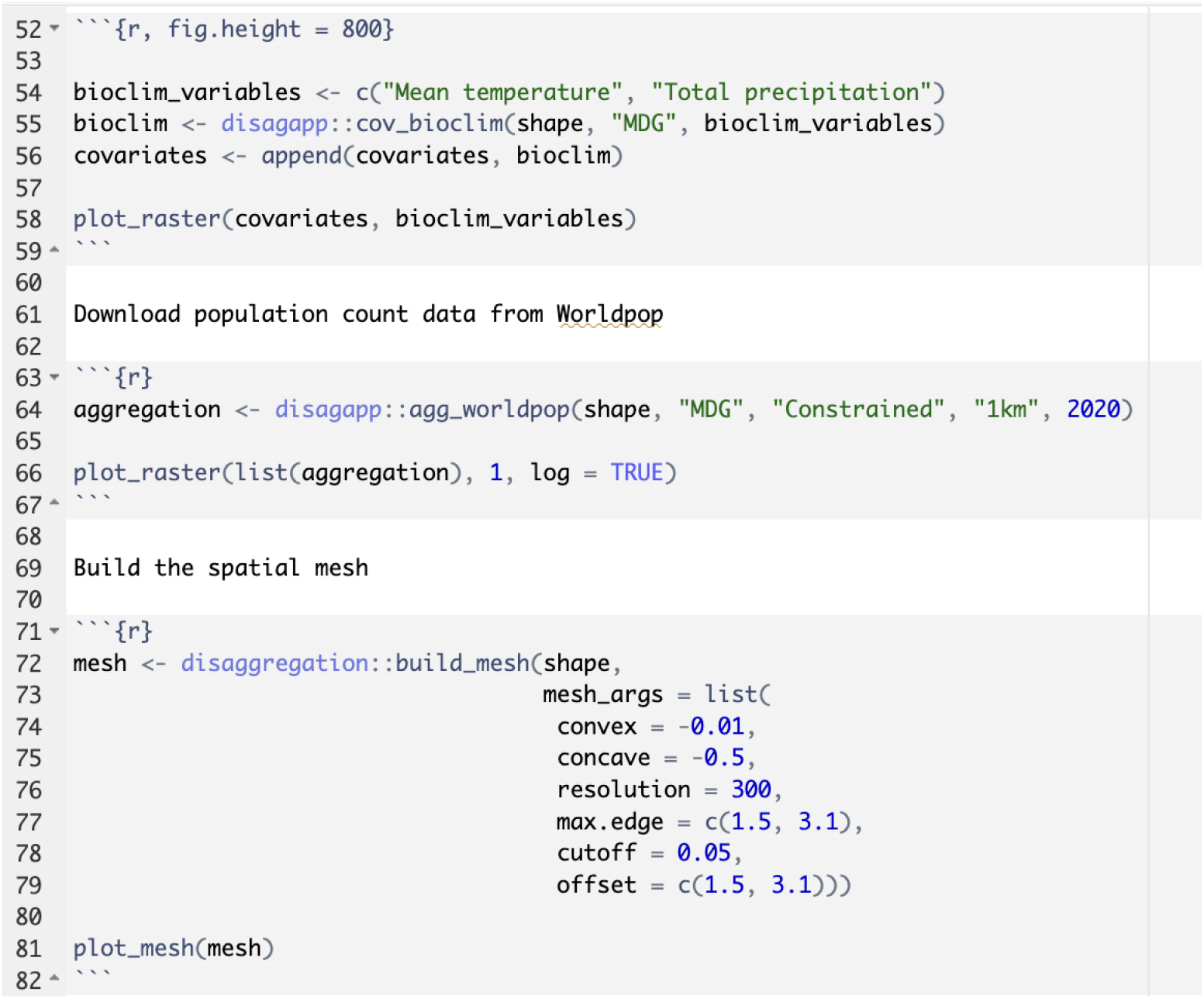
Screenshot of the Rmarkdown report generated by the *Session code* module after completing an analysis.

### 5.6 Saving and loading

The current state of the app can be saved at any point during an analysis and restored later, facilitating sharing between colleagues or storing incomplete analyses. To save the app, navigate to the *Save* tab in the results panel and click *Save Session* to download an `.rds` file. To restore the session at a later date, navigate to the *Load* tab in the *Intro* tab visible when the app is opened and upload the `.rds` file.

## 6 Conclusions

A web app has been developed that enables any researcher to conduct a disaggregation regression analysis through a web browser. By providing simple access to covariates and the modelling technique, we make this method much more accessible to the global modelling community. The app was designed to be easy-to-use and features practical and theoretical guidance on how to use it. Analyses conducted can be reproduced outside of the app by downloading an Rmarkdown document, supporting open science principles.

## Data Availability

No data were produced in this work. The source code is available online at: https://github.com/simon-smart88/disagapp

## 7 Availability and requirements

- Project name: Disagapp
- Project home page: https://simon-smart88.github.io/disagapp/
- Archived version: https://zenodo.org/records/17121965
- Operating system(s): Platform independent
- Programming language: R
- Other requirements: GDAL
- License: GNU GPL-3.0
- Any restrictions to use by non-academics: None

## 8 Declarations

### 8.1 Ethics approval and consent to participate

Not applicable.

### 8.2 Consent for publication

Not applicable.

### 8.3 Availability of data and materials

The source code is available from GitHub or Zenodo. The dataset analysed in the manuscript is available in the package.

### 8.6 Authors’ contributions

SS developed the package and prepared the manuscript. OO conducted user testing and provided feedback on usability. TL conceived the project, secured funding, recognised the potential to reuse Wallace functionality and supervised SS and OO. All authors reviewed the manuscript.

## 8.7 Acknowledgements

Not applicable.

